# Single-cell immune profiling at time of *M. tuberculosis* exposure reveals antigen-reactive programs that predict progression to active disease

**DOI:** 10.1101/2025.04.29.25326433

**Authors:** Jonathan Penã Avila, Joshua Simmons, Marina C. Figueiredo, Megan Turner, Marcelo Cordeiro-Santos, Valeria C. Rolla, Afranio L. Kristki, Rama Gangula, Cynthia Nochowicz, Ramesh Ram, Samuel Bailin, Simon Mallal, Silvana Gaudieri, Eric Alves, Beatriz Barreto Barreto-Duarte, Artur T. L. Queiroz, Helder I. Nakaya, Bruno B. Andrade, Timothy R. Sterling, Spyros A. Kalams, RePORT-Brazil Consortium

## Abstract

Early delineation of host immune responses at the moment of *Mycobacterium tuberculosis* (Mtb) exposure and infection is critical to identify individuals at risk of progressing to active tuberculosis (TB). We performed single-cell transcriptional profiling of over 500,000 peripheral blood mononuclear cells from 57 HIV-negative close contacts of TB cases in Brazil, including 25 individuals who developed active disease within two years (progressors) and 32 matched controls who remained disease-free (non-progressors). Cells were stimulated separately with the MTB300 peptide pool or irradiated Mtb (gRV), enabling resolution of antigen-reactive states across adaptive (CD4⁺ T-cells expressing abundant cytokines including IFNG, TNF, and IL17F) and trained-innate lineages, such as NK cells (producing GM-CSF, IFNG, CCL3, CCL4) and monocytes (GM-CSF, IL12B, IL36G). Progressors exhibited early hyper-metabolic CD4⁺ T-cell programs and proliferative NK cell signatures, whereas non-progressors preferentially upregulated complement activation and CCL3/4-driven chemokine signaling in monocytes. Notably, among progressors, gene expression profiles within antigen-reactive CD4⁺ T-cells and monocytes predicted the timing of progression to active TB. Together, these findings reveal high frequencies and functional diversity of antigen-reactive cells in Mtb-exposed individuals and nominate tractable immune correlates for the rational design of next-generation TB vaccines.

## Introduction

In most individuals, *Mycobacterium tuberculosis* (Mtb) infection leads to pulmonary granuloma formation, which controls or eliminates Mtb ^1, 2^. Both innate and adaptive cellular immune responses appear to be responsible for the control of Mtb infection. T cells are critical for successful containment of Mtb infection. Polyfunctional CD4+ T cells secreting IFN-γ, TNF-α and IL-2 are associated with the control of infection ^3^ and help macrophages control intracellular mycobacteria through the secretion of cytokines and cytotoxic T lymphocyte function ^4, 5^. Natural killer (NK) cells are innate lymphocytes that secrete IFN-γ and perform cytolytic functions to mediate the control of various pathogens, including Mtb. NK cells can mediate direct killing of Mtb-infected macrophages ^6^ but can also restrict intracellular bacterial replication via secretion of IL-22 ^7^ and IFN-γ ^8^. A population of IL-21-dependent NK cells that appear following BCG vaccination has been shown to expand following Mtb challenge ^9^, suggesting that NK cells may also display some hallmark characteristics of memory cells ^10^. While disruption of control of chronic infection can be a consequence of immune suppression, either from HIV infection ^11^, or from medications such as TNFa inhibititors or corticosteroids ^12^, other individuals progress to active infection without obvious inciting reasons.

Identification of Mtb-reactive cells typically relies on cytokine production by T cells, as measured by ELISPOT, intracellular cytokine staining, or cytokine measurement in the supernatant after in vitro stimulation of cells with Mtb antigen or peptides ^13, 14^. Diagnostic interferon release assays (IGRAs) rely on the specificity of the immune response to Mtb antigens such as ESAT-6, CFP10, and TB7.7 and are used to provide evidence of Mtb infection ^2, 13, 15, 16, 17, 18, 19, 20, 21, 22, 23^. More recent studies have identified Mtb antigen-specific T cells by the expression of surface activation markers ^24, 25^. Compared to that of cytokine expression, description of the transcriptional profile of Mtb-reactive cells has been limited. Mtb-reactive CCR6+CXCR3+CCR4-cells are enriched in Mtb-infected individuals. After bulk sorting and stimulation with PMA/ionomycin they display a transcriptional profile with features of Th1 and Th17 cells, with enrichment of genes related to cytokine/receptor interactions ^26^. At the single-cell level, transcriptional profiling has been used to differentiate Mtb infection from active TB disease without identifying antigen-specific cells ^27^.

In this study, we comprehensively analyzed Mtb antigen-reactive cells at the single-cell level among close TB contacts. This was performed on cells obtained after TB exposure, included IGRA-positive and -negative persons, and individuals who subsequently progressed to active TB vs. those who did not, over 2 years of follow-up. Peripheral blood mononuclear cells (PBMCs) were stimulated overnight separately with irradiated Mtb organisms or a “megapool” of 300 optimal Mtb peptide epitopes ^28^. Our goal was to identify populations of Mtb-reactive immune cells and transcriptional pathways associated with progression to active disease or control of infection. These studies provide new insights into Mtb pathogenesis, will help identify individuals at the highest risk of progression to TB, and will potentially identify immune correlates of protection elicited by Mtb candidate vaccines.

## Results

### Study cohort description

All study participants were from the Regional Prospective Observational Research in Tuberculosis (RePORT)-Brazil cohort ^29^. For this study, 58 HIV-seronegative close contacts of individuals with culture-confirmed pulmonary TB were selected; all were asymptomatic and had no radiographic evidence of active TB at enrollment. One participant had insufficient cells and was thus excluded. Of the 58 close TB contacts, 25 subsequently progressed to active, symptomatic TB within 24 months of follow-up; this represented all TB progressors in RePORT-Brazil. There were 33 controls who were also close TB contacts, but did not progress to TB within 24 months. The controls were matched to progressors based on age (+/- 5 years) and sex; they received either no therapy or < 30 days of isoniazid preventive therapy. All 58 close TB contacts were HIV-seronegative. The clinical and demographic characteristics of the study population are shown in **Table 1**. There was no statistically significant difference according to progressor status with regard to age, sex, self-reported race, body mass index (BMI), or BCG vaccination status. Moreover, there were no significant differences in source case cavity according to chest X-ray, AFB smear positivity, tobacco use, or dysglycemia (**Table 1**).

**Table 1.** Clinical and demographic characteristics at baseline of the study cohort of close TB contacts according to progressor status (n = 57).

| Characteristic | Total (n=58) | Progressor<br>n=25 | Control<br>n=33 | P-value |
| --- | --- | --- | --- | --- |
| Female sex | 38 (66%) | 16 (64%) | 22 (67%) | 1.0 |
| Age, median (IQR) | 34.0 (18.5-50.9) | 31.9 (18.3-50.1) | 35.4 (19.2-52.4) | 0.83 |
| Race, self-reported |  |  |  | 0.73 |
| White | 12 (21%) | 6 (24%) | 6 (18%) |  |
| Black | 20 (34%) | 7 (28%) | 13 (39%) |  |
| Brown/pardo | 25 (43%) | 12 (48%) | 13 (39%) |  |
| Indio | 1 (2%) | 0 (0%) | 1 (3%) |  |
| BMI median (IQR) | 24.5 (21.7-28.7) | 24.2 (19.6-25.6) | 25.3 (21.9-30.7) | 0.08 |
| BCG vaccine scar | 51 (88%) | 21 (84%) | 30 (91%) | 0.45 |
| Index case CXR cavity – yes | 38 (66%) | 15 (60%) | 23 (70%) | 0.58 |
| Index case smear + AFB | 46 (79%) | 20 (80%) | 26 (79%) | 1.0 |
| Index case tobacco use |  |  |  | 0.79 |
| Current | 12 (21%) | 4 (16%) | 8 (24%) |  |
| Former | 20 (34%) | 9 (36%) | 11 (33%) |  |
| Never | 26 (45%) | 12 (48%) | 14 (42%) |  |
| Index case DM | 15 (26%) | 7 (28%) | 8 (24%) | 0.7 |
| Index case pre-DM | 20 (34%) | 7 (28%) | 13 (39%) |  |
| Index case normal glycemia | 23 (40%) | 11 (44%) | 12 (36%) |  |

**Abbreviations**
IQR: interquartile range
BMI: body mass index
BCG: bacille Calmette-Guerin
CXR: chest X-ray
AFB: acid-fast bacilli
DM: diabetes mellitus; defined as HbA1c > 6.5
Pre DM: pre-diabetes mellitus; defined as HbA1c 5.7 – 6.5
Normal glycemia: defined as HbA1c < 5.7

### Identification of major cell populations

Cells were incubated overnight in media alone, with MTB300 megapool, or with gamma-irradiated *M. tuberculosis* strain H37Rv (gRV) and subjected to single-cell RNAseq. RNA sequence data were clustered to identify major cell lineages. Cells with evidence of activation (labeled as “antigen-reactive” cells) were further characterized at the transcriptional level to identify differences between progressors and non-progressors (**Figure 1**). An overlay of antibody-derived tags (ADTs) from a CITE-seq panel confirmed lineages such as T cells (CD3+, CD4+, CD8+)(**Supp. Fig. 1 a,b**), NK cells (CD16+, CD56+) (**Supp. Fig. 1 c,d**), monocytes and dendritic cells (CD11c)(**Supp. Fig. 1 e**) and B cells (CD19)(**Supp. Fig. 1 f**). (Having identified major immune cell populations, we next performed a more detailed analysis of the T, NK, amd monocyte antigen-reactive cell populations.

**Figure 1.**
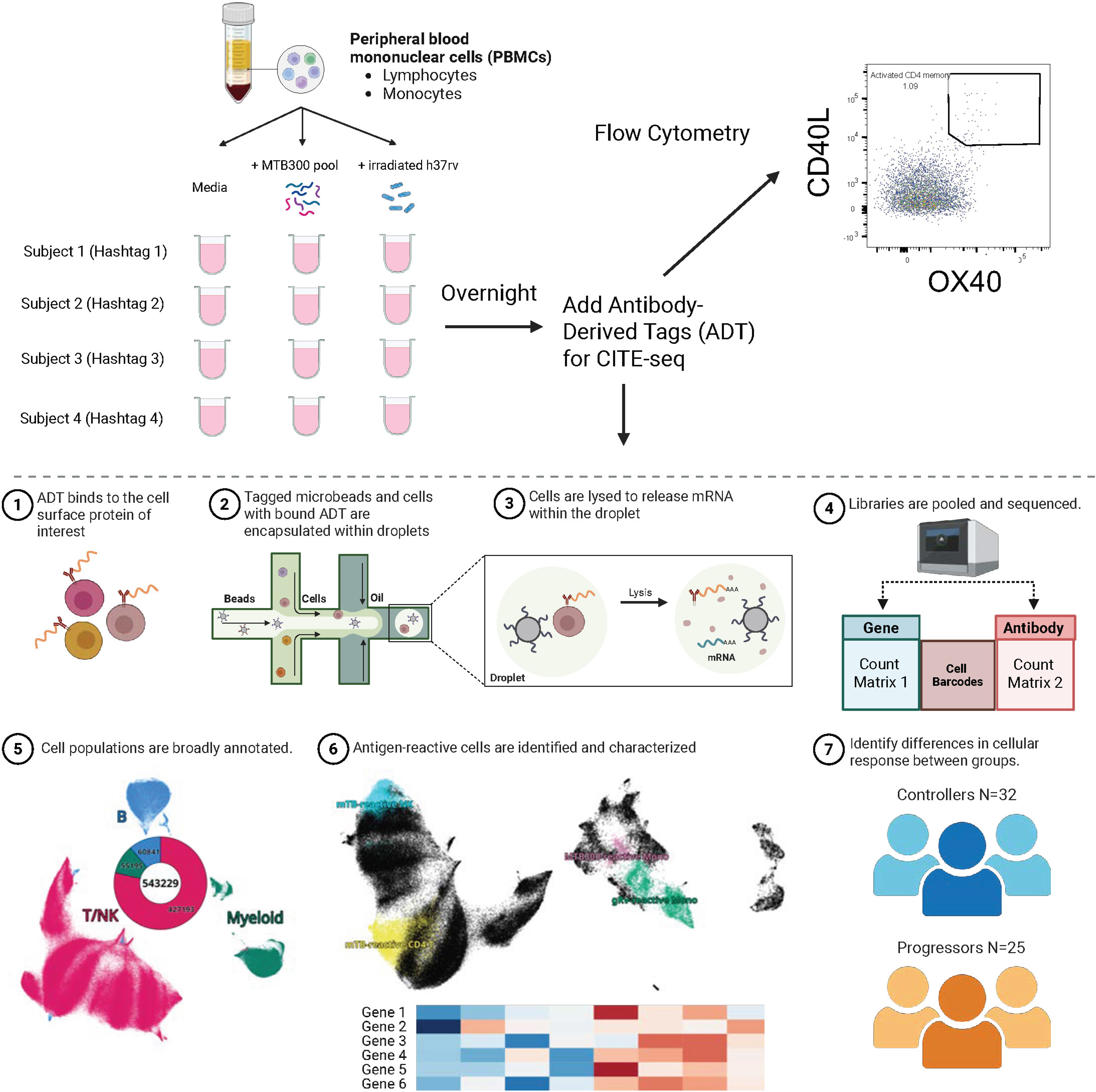
10X and CITE-seq workflow. After overnight incubation with MTB300 or gRV, antibody-derived tags (ADT) were added, and cells were subjected to scRNAseq analysis. A portion of cells were incubated with antibodies for flow cytometry analysis.

### Transcriptomic profile and surface protein expression of antigen-reactive T and NK cell populations

Compared to those in the media (control), several cell cluster frequencies were significantly higher in the presence of Mtb antigens. After exporting, and re-integrating T and NK cell populations, we used scCODA^30^ to identify cell populations that changed with frequency in the presence of antigen. In **Figure 2A**, we show annotated cell populations identified by a combination of gene expression profile and surface protein expression. Using CD4+ T cell naïve cells as a reference population, which was unchanged in frequency in the presence of antigen, scCODA identified a subset of CD4+ T cells tat credibly declined in frequency (CD4 T memory cluster 1, **Figure 2C**), and 3 populations that credibly increased in frequency in the presence of either MTB300 or gRV (cell clusters 9, 21, and 27, **Fig 2 D-F**). A similar analysis of NK cells identified cell cluster 10 as credibly decreasing in the presence of antigen (**Supplemental Figure 2)**, and Mtb-reactive NK cells increasing in abundance in the presence of either MTB300 or gRV (**Fig 2G**). The transcriptional profile and surface marker expression of antigen-reactive CD4+ T cells were consistent with T cell activation. Each of the identified cell populations (cell clusters 9, 21, and 27) had different degrees of expression of activation genes such as TNFRSF4 (OX40), TNFRSF9 (CD137), CD25 (IL2RA), and CD69, chemokines (CCL3, CCL4, CSF2), and cytokines (IFNG, IL2, TNFa, IL17F, IL22, IL32) (**Fig 2H**). The corresponding profiles of these cells by surface protein expression was consistent with the transcriptional profiles, and included activation markers such as CD25, CD69, CD137, and KLRG1 (**Fig 2I**). Of these 3 activated CD4+ T cell populations, cluster 27 had the highest degree of immune activation, but as they were highly similar to each other they were grouped together for downstream analysis of differences between progressors and non-progressors. They are shown as Mtb-reactive CD4 T cells in **Figure 2 H-I**. Activated Mtb-reactive NK cells showed a transcriptional profile similar to activated T cells, with high transcript expression of cytokines (IFNG, TNF), chemokines (CCL3, CCL4) and activation markers (CD69, TNFRSF4, TNFRSF9) and corresponding surface expression of activation markers CD25, CD69, CD137 (**Fig 2 J-K**).

**Figure 2.**
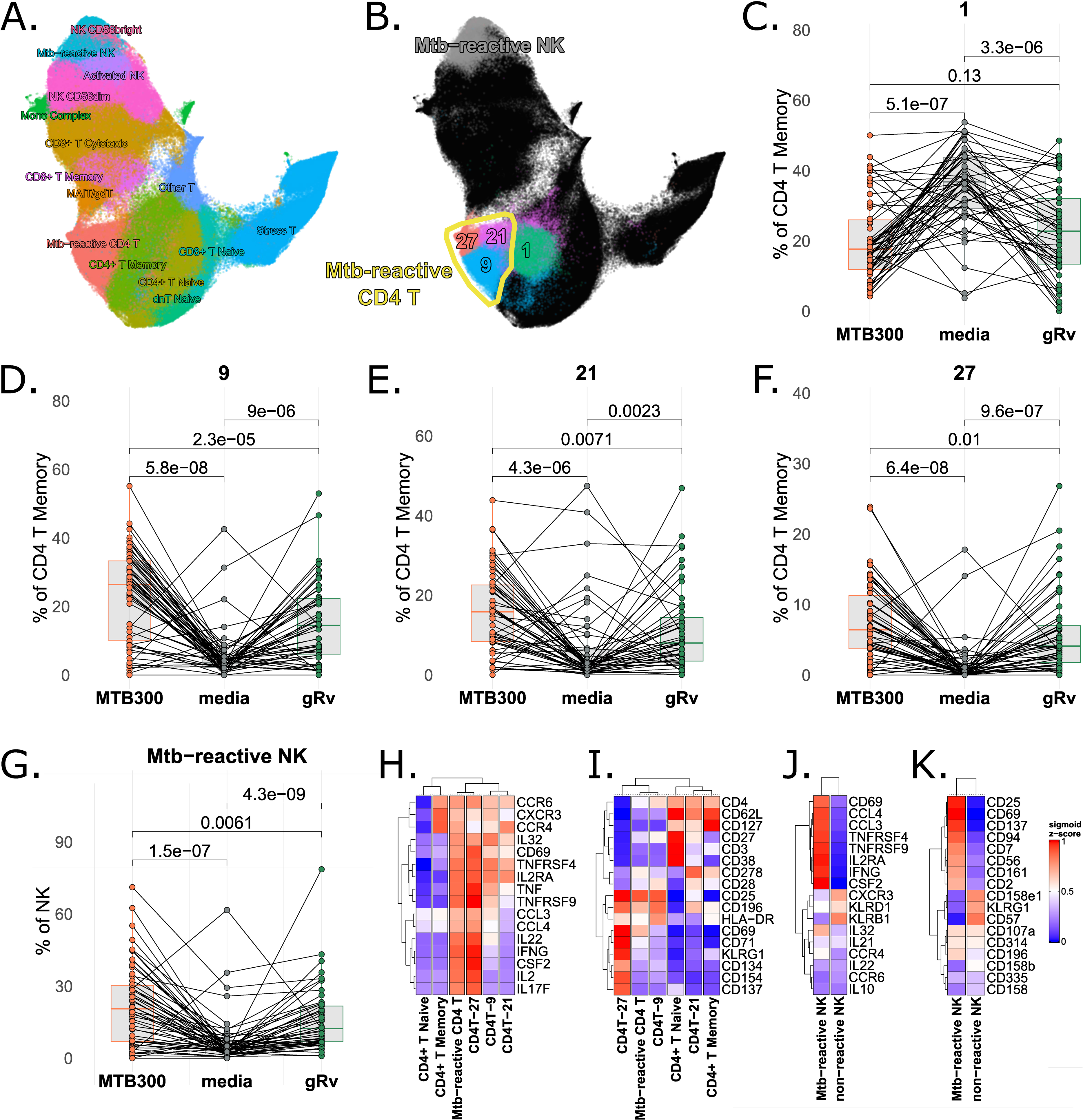
Abundance, transcriptomic profile and surface protein expression of T and NK cells. A. Cell population annotations of T and NK cells. B. UMAP and annotations of T/NK cell populations that change in abundance in response to antigen stimulation. C-G. Frequencies of cell populations as a percentage of T/NK cells after overnight antigen stimulation. C. Cell cluster 1; central memory CD4+ T cells. D. CD4+ T cell cluster 9. E. CD4+ T cell cluster 21. F. CD4+ T cell cluster 27. G. Mtb-reactive NK cells. H. Transcriptional profile of CD4+ T cells. I. ADT expression of CD4+ T cells. J. Transcriptional profile of NK cells. K. ADT expression of NK cells.

### Monocyte populations show distinct transcriptomic profiles depending on antigen stimulation

Monocytes were classified by combinations of gene expression and surface markers (**Fig 3A**). We found 3 populations of monocytes that preferentially increased in frequency in response to the gRV antigen peptide pool compared to MTB300 (cell clusters 0, 6, 8; **Fig 2 C-E**). These cell clusters were transcriptionally similar, expressing high levels of CSF2, IL36B, CCL2 and IL12B, and were grouped together as gRV-reactive monocytes for further analysis (**Fig 2G**, **Fig 3G**). A separate population of monocytes preferentially responded to MTB300 and expessed high levels of HLA Class II molecules (MTB300-reactive mono; **Fig 2H**, **Fig 3G**).

**Figure 3.**
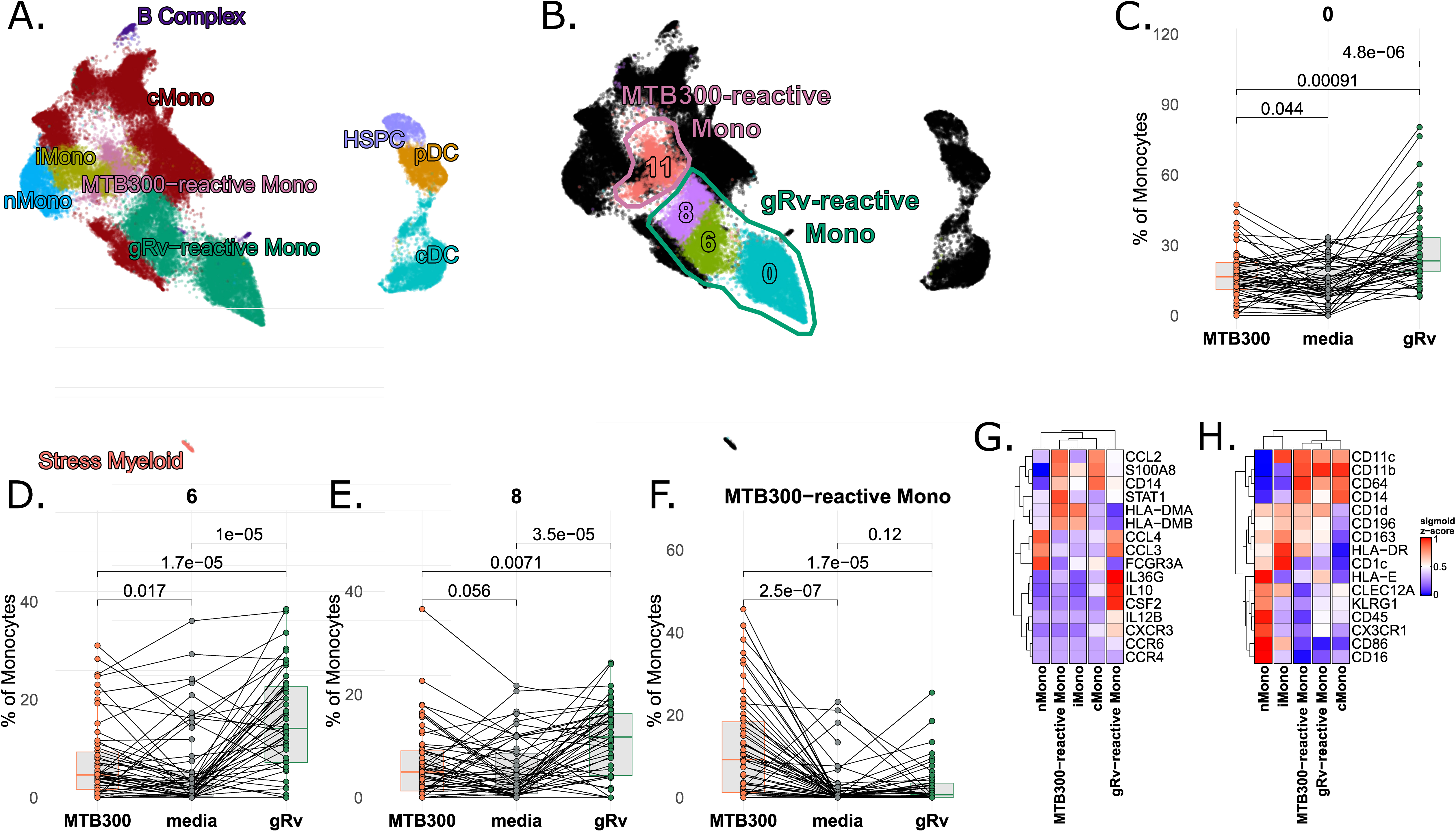
Abundance, transcriptomic profile and surface protein expression of myeloid cells. A. Cell population annotations of myeloid cells. B. UMAP of Cell populations that change in abundance in response to antigen stimulation. C-F. Frequencies of cell populations as a percentage of myeloid cells after overnight antigen stimulation. C. gRv-reactive monocyte cell cluster 0. D. gRv-reactive cell monocyte cluster 6. E. gRv-reactive monocyte cell cluster 8. F. MTB300-reactive monocytes. G. Transcriptional profile of MTB300-reactive monocytes. I. ADT expression of MTB300-reactive monocytes. J. Transcriptional profile of gRv-reactive monocytes. K. ADT expression of gRv-reactive monocytes.

### Myeloid and CD4+ T cell populations show distinct transcriptional patterns in progressors and non-progressors

Our goal was to characterize differences in antigen responsiveness between groups based on progressor status. There was no significant difference in the frequency of either MTB300-reactive mono nor gRv-reactive mono between progressors and non-progressors (**Supp. Fig 3 A, B**). Likewise, we found no differences in frequencies of Mtb-reactive CD4+ T cells between groups after stimulation with MTB300 or gRv (**Supp. Fig 3C**). Frequencies of NK-reactive cells were higher under stimulation conditions in progressors compared to non-progressors, but only the frequency difference in the presence of gRv was significant (**Supp. Fig 3D**; p=0.023). We next focused on transcriptional differences between these reactive cell populations in progressors compared to non-progressors.

In **Figure 4** we show the results of gene set enrichment analysis (GSEA) after gRv stimulation of PBMC. In **Figure 4A** we show that gRv-reactive monocytes in non-progressors had pathways enriched for protein translation (gene set cluster 1), immune activation (creation of C3 and C2 activators and FCGR activation; gene set cluster 2), chemokine signaling (peptide ligand binding receptors and chemokine receptors bind chemokines; gene set cluster 4) and PD-1 signaling (gene set cluster 6). Conversely, gRV-reactive CD4+ T cells in progressors had enriched pathways for protein translation (gene set cluster 1), as well as metabolic pathways such as respiratory electron transport and aerobic respiration (gene set cluster 7), mitochondrial translation (gene set cluster 9), mRNA processing (gene set cluster 10), and other protein processing pathways (gene set cluster 11). In **Figure 4B** we show the enrichment plot for the reactome chemokine receptors bind chemokines gene set in gRv-reactive monocytes and the reactome mRNA splicing pathway in Mtb-reactive T cells. In **Figure 4C** we show distribution of leading edge gene scores for two of the gene sets for the cell types in which they were found to be significant, stratified by progression status. Scores were calculated by taking the average z-score of the edgeR log2 CPM counts for the leading edge genes.

**Figure 4.**
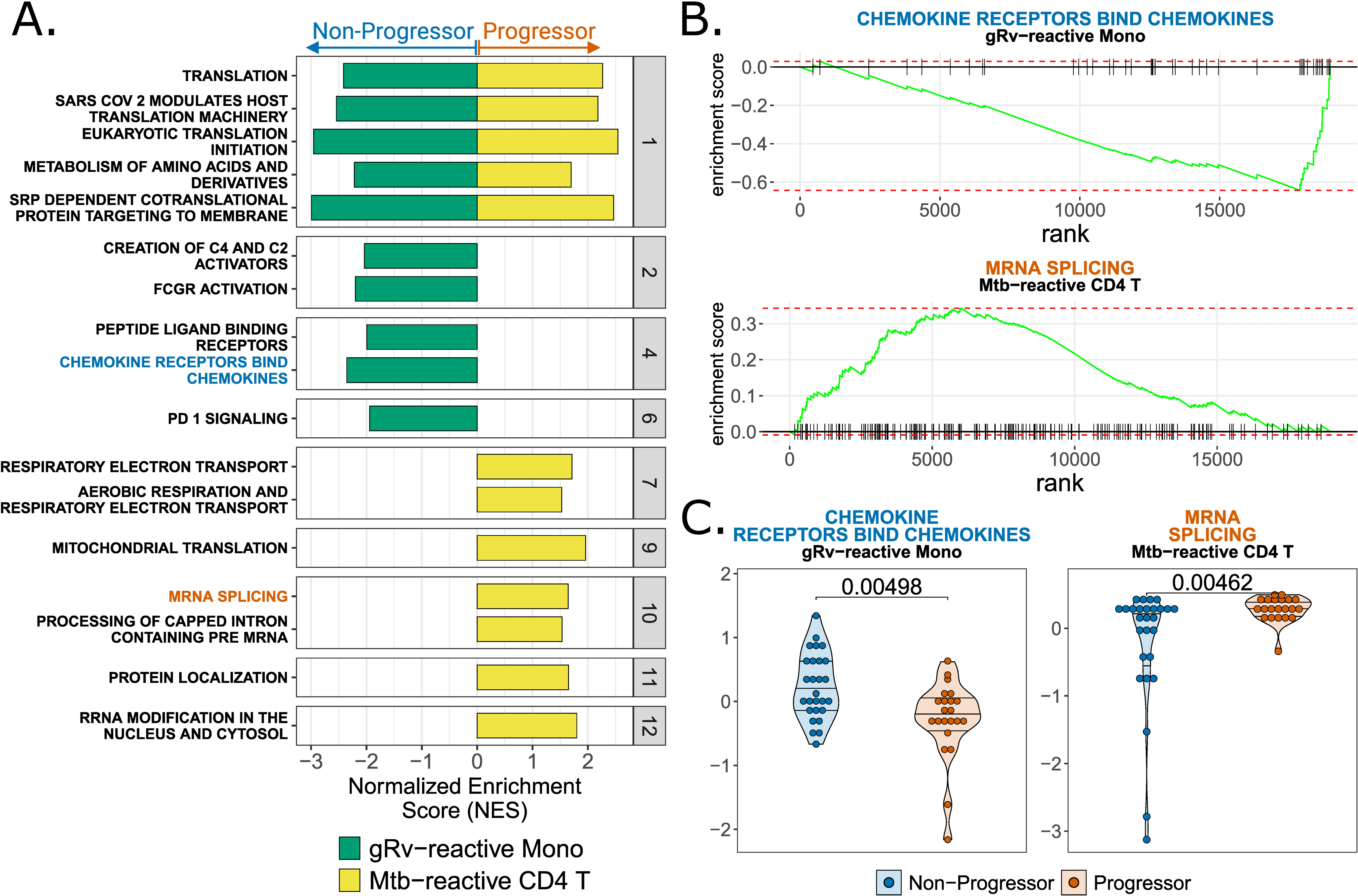
TB non-progressors and progressors show distinct transcriptional profiles of Mtb-reactive cells after gRv stimulation. **A.** Gene set enrichment showing differences in gRv-reactive monocytes and Mtb-reactive CD4+ T cells in non-progressors compared to progressors. **B**. Enrichment plots of representative pathways, reactome chemokine receptors and chemokines in gRv-reactive monocytes and reactome mRNA splicing in Mtb-reactive CD4+ T cells. **C.** Leading edge score differences between non-progressors and progressors for each gene set in (**B**).

We found similar differences between non-progressors and progressors after MTB300 stimulation. In response to peptide stimulation, MTB300-reactive monocytes in non-progressors had a transcriptional pattern consistent with immune activation (FCGR activation), protein translation (eukaryotic translation elongation) and production of chemokine and chemokine receptors (peptide ligand binding receptors). Mtb-reactive NK cells showed pathways upregulated in progressors, including those related to cell replication (cell cycle checkpoints, DNA replication, DNA strand elongation), interferon signaling (interferon alpha beta signaling, interon gamma signaling) and antigen presentation (MHC Class II antigen presentation)(**Figure 5A**). In **Figure 5B** we show show distribution of leading edge gene scores for reactome peptide binding receptors, which are higher in MTB300-reactive mono in non-progressors (**Figure 5C**, p=0.0007), and MHC Class II antigen presentation genes, which are higher in Mtb-reactive NK cells in progressors (**Figure 5C**, p=0.0008). Overall, in response to antigen stimulation, MTB300-reactive monocytes in non-progressors showed transcriptional programming associated with protein translation, antigen presentation, and chemokine signalling. In progressors, the primary transcriptional programs directed metabolic activity, increased protein translation, cell proliferation, and cytokine signaling.

**Figure 5.**
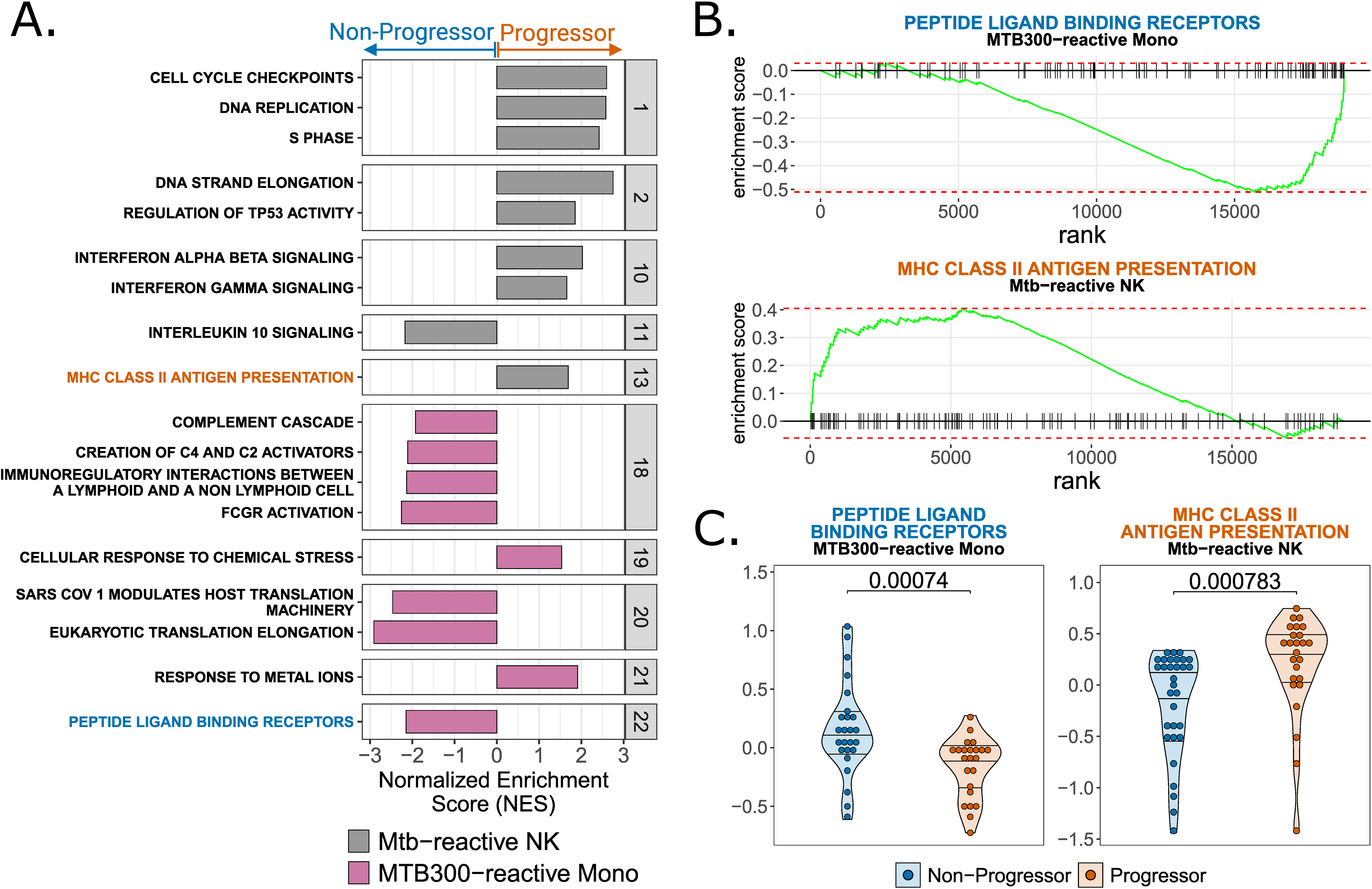
TB non-progressors and progressors show distinct transcriptional profiles of Mtb-reactive cells after MTB300 stimulation. **A**. Gene set enrichment showing differences in MTB300-reactive monocytes and Mtb-reactive NK cells in non-progressors compared to progressors. **B**. Enrichment plots of representative pathways, reactome peptide ligand binding receptors in Mtb-reactive monocytes and reactome MHC Class II antigen presentation Mtb-reactive NK cells. **C**. Leading edge score differences between non-progressors and progressors for each gene set in (**B**).

### Gene networks demonstrate related pathways, and genes predicting progression vs. control in antigen-reactive innate and adaptive immune cell populations

We performed a network analysis of enriched pathways to help visualize similarities and differences in pathways between antigen-reactive cell clusters at the gene level. In **Figure 6A** we show the networks of the top 30% ranked genes for gRv-reactive mono. Networks associated with control of infection included chemokine signaing (several chemokines and chemokine receptors) and translation. We observed a similar pattern with MTB300-reactive monocytes (**Figure 6B**), with chemokine and chemokine receptor gene expression, and gene expression of markers signifying interactions with NK cells (KLRB1 and KLRD1) associated with control of disease. In this monocyte population there were also networks associated with disease progression, including several MT1 genes that regulate zinc transport, and HB genes, indicating a response to stress. Mtb-reactive NK cells in progressors showed higher expression of genes associated with replication (cell cycle checkpoints) and class II antigen presentation.

**Figure 6.**
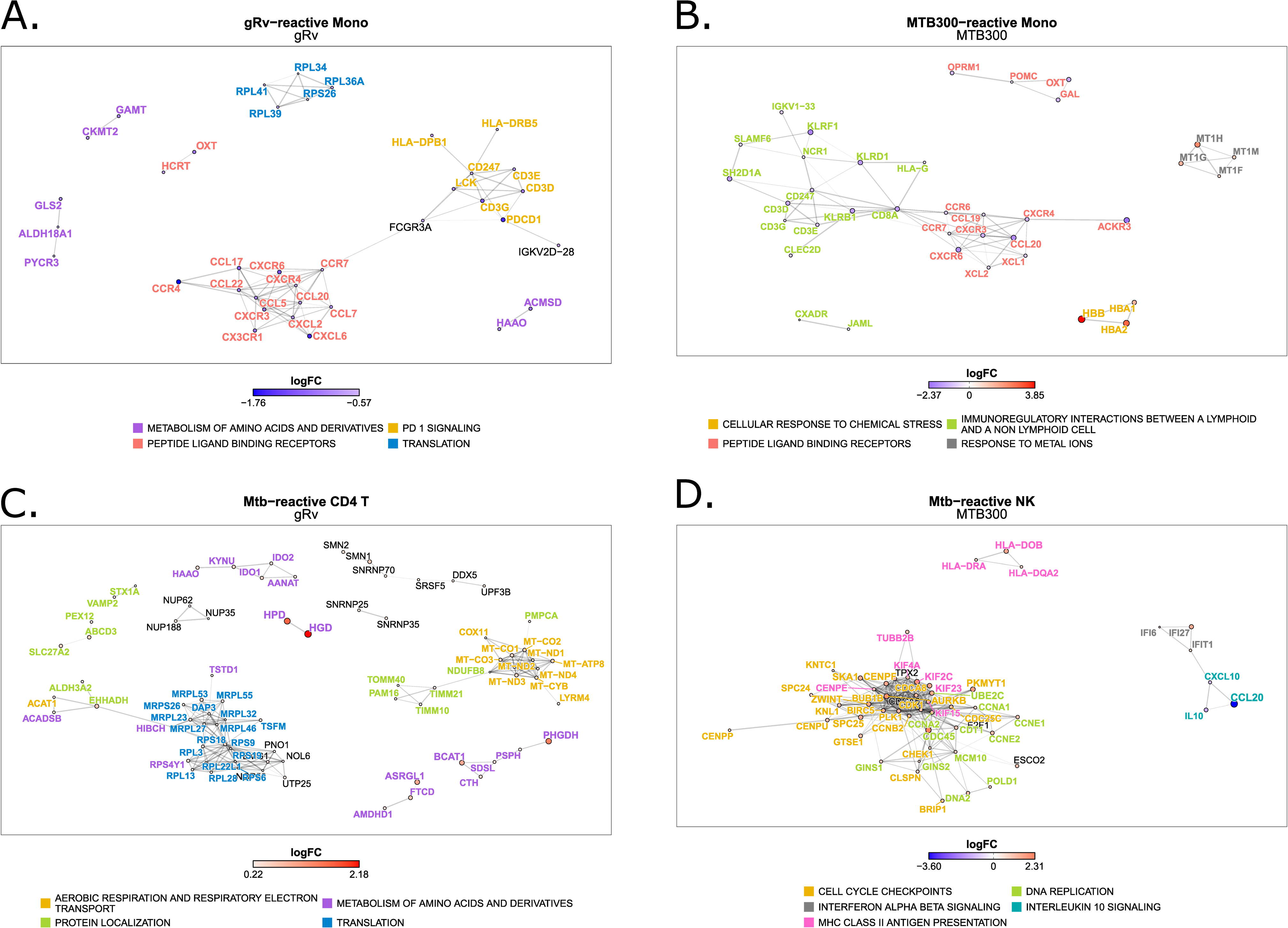
Gene-gene STRING protein-protein interactions of gene networks associated with control or progression of TB. A. Gene networks of monocytes reactive to gRv (gRV-Mono). B.Gene networks of monocytes reactive to MTB300 (MTB300-reactive Mono). C. NK cells reactive to MTB300 (Mtb-reactive NK). D. CD4+ T cells reactive to gRV (Mtb-reactive CD4). Color scale indicates log fold change of gene expression in each cell population, blue toward control, red toward progression.

Genes associated with control included IL-10 and interferon alpha/beta signaling (**Figure 6C**). gRv reactive CD4+ T genes associated with progression were associated with cellular activation, protein translation, and metabolism (electron transport) (**Figure 6D**).

### Gene expression profiles of antigen-reactive CD4+ T cells predict time to disease progression

Having found several gene expression profiles predicting progression to or control of disease, we next evaluated whether the response of cells to antigen could predict the pace of progression to disease in TB progressors. In **Figure 7** we show gene network scores that predicted progression in Mtb-reactive CD4 T cells after stimulation with gRv. These profiles are associated with cell replication and metabolism, including electron transport (**Figure 7 A, D**), mitochondrial protein degradation, protein translation (**Figure 7B**) and mitochondrial protein degradation (**Figure 7C**).

**Figure 7.**
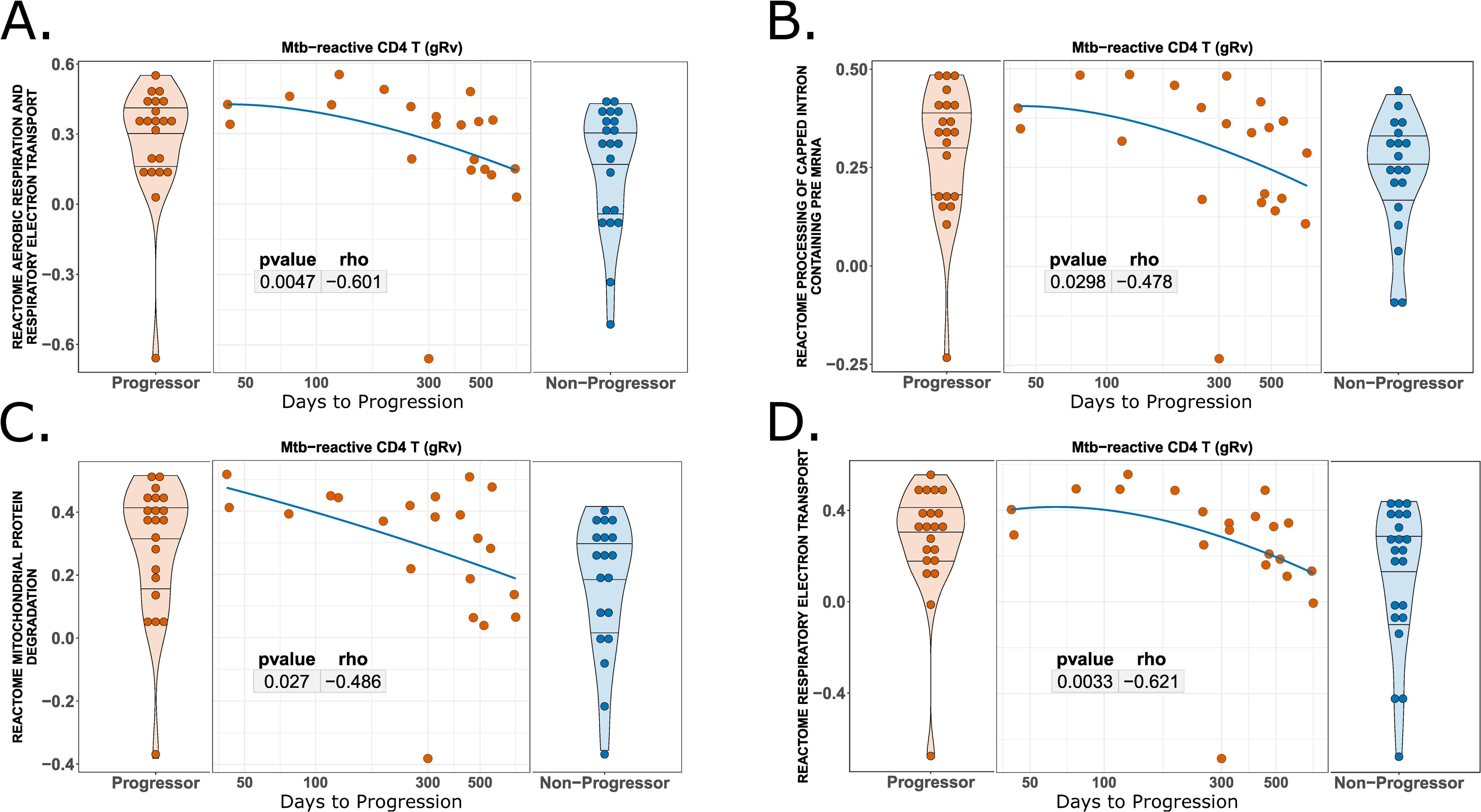
Gene set scores predicting progression to active TB in gRv-reactive CD4 T cells. A. Aerobic respiration and electron transport B. Processing of capped intron containing pre-RNA. C. Mitochondrial protein degradation. D. Respiratory electron transport.

## Discussion

We identified profiles of Mtb-reactive innate and adaptive immune cells that predicted progression to active TB or control of infection in a cohort of individuals exposed to Mtb, but without evidence of active disease at baseline. We identified several populations of reactive cells which are thought to play a role in maintaining control of Mtb infection.

We found 2 discrete populations of Mtb-reactive monocytes, depending on whether cells were stimulated with gRv or with the MTB300 peptide pool. The profiles were similar, but in response to gRv, monocytes had a transcriptional profile associated with chemokine and chemokine receptor expression, whereas in response to peptides, the profile had high expression of HLA class II genes. However, both populations of cells showed gene expression patterns governing protein translation and chemokine signaling that were higher in non-progressors compared to progressors.

We found significant increases in NK cell activation in response to Mtb antigen stimulation, which is part of trained immunity. NK cells augment immunity against Mtb indirectly by enhancing CD8 T-cell production of IFN-γ ^31^, by promoting the expansion of γ8 T-cells ^32^, and by lysing Mtb-expanded regulatory T-cells ^33^. Other studies have identified that CD25^+^/CD54^+^/CD94^+^ NK cells represent a highly specialized subpopulation with improved survival, elevated IFNγ secretion and cytolytic activity, particularly following IL-15 exposure.^34, 35, 36^ We found higher frequencies of NK-cells after in vitro stimulation with gRv in progressors compared to non-progressors. The gene profiles of these cells in progressors was characterized by gene networks associated with cell division, as well as increased expression of interferon alpha/beta and interferon gamma signaling genes.

Both groups had high frequencies of antigen-reactive CD4+ T cells, and the frequency of cells was not significant between progressors and non-progressors. CD4+ T cells provide important helper functions for other immune cells involved in the response to Mtb infection, including facilitating the responses of CD8+ T cells, and antibody production by B cells ^2, 37^. Detailed analyses of immune responses to Mtb proteins and peptides have revealed a broad range of immune recognition across multiple Mtb proteins, primarily involving HLA class II-restricted CD4+ T-cell memory responses with combinations of IFNγ, TNFα, and IL-2 cytokine secretion, and phenotypically dominated by a CCR6+CXCR3+CCR4-chemokine receptor expression profile ^38, 39, 40^.

Mtb antigen-reactive CD4+ T cells had high expression of activation markers such as TNFRSF4 (OX40), TNFRSF9 (CD137) and IL2RA (CD25). Cytokine transcripts included IL2, IFNG, IL22, and IL17F, LTA, LTB, and IL32. LTA and LTB are members of the TNF superfamily ^41, 42^ and play an important role in the control of intracellular pathogens such as *Listeria monocytogenes* and Mtb ^43^. IL32 is produced by several cell types, including T cells and NK cells. Early studies revealed that Mtb antigen potently induces IL-32 expression in PBMCs ^44^. IL-32 induces TNFα production in PBMCs ^45^ and transforms monocytes into macrophage-like cells with increased phagocytic ability ^46^. Similar to the profiles in NK cells, Mtb-reactive CD4 T cells in progressors had higher expression of genes associated with translation and electron transport.

In addition to gene programs that differentiated progressors and non-progressors, we also showed that among progressors, some of these programs were related to the time to progression. In the case of antigen-reactive CD4+ T cells, individuals with a shorter time to progression showed gene expression profiles consistent with increased metabolic activity (mitochondrial electron transport and mitochondrial protein degradation) and mRNA processing.

The nature of this study did not allow us to isolate direct responses of cells to antigen, but rather reflected how cells in culture responded both directly to antigen and each other through direct contact and cytokine signalling pathways. Future studies can refine these interactions by either depleting and adding specific cell subsets, or by targeting some of the genes we have identified.

Our specific goal in this study was to identify and describe populations of innate and adaptive Mtb antigen-reactive cells at the transcriptional level, and to identify gene expression profiles associated with progression to TB or control of Mtb infection. The cell populations we identifed were likely to play synergistic roles in the maintenance of recognition and control of Mtb replication after infection ^10, 47, 48^. We demonstrated that the relative abundance of antigen-reactive cells did not differ greatly between progressors and non-progressors, yet were differentiated at the transcriptional level. Analysis of these cell populations and their interactions with antigen-reactive cells will provide insight into the ability of individuals to maintain long-term control of Mtb infection, uncover networks of responses elicited after successful vaccine strategies, as well as identify persons at high risk of progression to TB.

## Methods

### Study participants

Study participants were from the Regional Prospective Observational Research in Tuberculosis (RePORT) Brazil cohort ^29^. For this study, we included persons who were close contacts of individuals with culture-confirmed pulmonary TB, regardless of interferon gamma release assay (IGRA) status.

### Antigen stimulation

Cryopreserved PBMCs were thawed in the presence of the nuclease S7 [Sigma Aldrich (Roche), St. Louis, MO]. The cells were counted, washed with R10, and resuspended at 10×10^6^ cells/ml. An anti-CD40 blocking antibody (Miltenyi Biotech, Auburn, CA) was added to the cells at a final concentration of 0.5 μg/ml (1:200 dilution) for 15 minutes. The cells were subsequently added to the wells of a 96-well plate (1 million cells per stimulation condition), and the anti-CD154 PE antibody was added to each well at a 1:10 final dilution. The cells were stimulated with media alone, MTB300 megapool (kindly provided by Cecilia Lindestam Arlehamn, final concentration of 2 μg/ml) or gamma-irradiated h37RV (10 μg/ml) (BEI Resources).

### Library preparation and sequencing

After overnight incubation, the cells from each well were washed with staining buffer, incubated with human true-stain FcX blocking reagent on ice for 10 minutes, and subsequently subjected to hashtag antibody staining for 30 minutes. The cells were washed 3 times with phosphate-buffered saline (PBS) supplemented with 2% bovine serum albumin (BSA) and centrifuged for 10 minutes at 400 × g between washes. The cells were then stained with a 137-antibody CITE-seq panel (Biolegend; Cat# 399905) on ice for 30 minutes and washed twice with PBS and 1% BSA ^49^. Each experimental run involved 4 or 5 individuals and consisted of 3 lanes for 10x sequencing (**Figure 1**). We targeted at least 20,000 cells per lane for each experimental run.

### Flow cytometry

After removing cells for downstream single-cell RNAseq analysis, the remaining cells in each well were counted and stained with the following antibody panel: anti-CD8 APC-Cy, -CD137 APC, -CD3 AF700, -CD45RO PE CF594, -OX40 PE-Cy7, -CD4 PcPCy5.5, -CD69 BV605, and -CD25 BV786 and LDaqua V500. CD4+ responses to each antigen were defined as %CD4+OX40+CD154+ and are displayed as a percentage of memory (CD45RO+CCR7+) CD4+ T cells (**Supplemental Figure 2**).

### Cellular indexing of transcriptomes and epitopes (CITE-seq)

Raw sequences (FASTQ files) were processed with 10x Genomics Cell Ranger Multi v9.0.0 using 10x Genomics Cloud Analysis ^50^. Reads were aligned to an amended GRCh38 reference with greater KIR gene resolution ^51^. The souporcell algorithm ^52^ was used for de-multiplexing of droplet barcodes and annotation of inter-sample doublets. Outlier cells by >5 median absolute deviations (MADs) for several QC factors and cells with high mitochondrial gene (MT-) counts (>10%) were removed. Souporcell genetic clustering from genomic sequencing of this data set ^53^ paired with hashtag count results were used to map sample ID to barcodes. Souporcell genotypic cluster SNP and genomic sequencing SNP correlation was quantified using the Demuxafy Assign_Indiv_by_Geno algorithm ^54^. The results were used for small adjustments to the barcode assignments, where appropriate. The Solo doublet-finding algorithm ^55^ implemented through the scVI python library ^56, 57^ was used to annotate intra-sample doublets. Inter- and intra-sample doublets were then removed. The final cell count was 543,229. Genes not expressed in at least 150 cells were removed and the final gene count was 18,968. Gene count imputation was applied to the raw concatenated count matrix using Adaptively-thresholded Low Rank Approximation (ALRA) ^58^. A consensus set of highly variable genes (HVG, n = 5000) were calculated on the imputed counts across project batches. Principal component analysis (PCA, n = 30) was performed on the HVG matrix. The Harmony algorithm ^59^ implemented through the harmonypy python library was used for integration of the PCA embedding with theta = 2 and batch ID set to patient sample sequencing group (n=13). Nearest neighbors (kNN, n = 30) were found using the Harmony components. Leiden clustering ^60^(resolution = 3) was used to partition the kNN graph. Cell clusters were split into three groups: T/NK cells, myeloid cells, and B cells based on cluster gene markers. Each subset was then analyzed separately by HVG, PCA (n = 30), Harmony, kNN (n = 30), and Leiden clustering.

Uniform Manifold Approximation and Projection (UMAP) ^61^was used for visualization. Leiden clusters were annotated according to marker expression profiles. Subset-specific HVG n-genes were set as follows: T/NK (n = 4000), myeloid (n = 2500), and B (n = 2000). Harmony theta was set as follows: T/NK (n = 4), myeloid (n = 4), and B (n = 2). Leiden resolution was set as follows: T/NK = 2.5, myeloid = 1, and B = 1.

The scCODA^30^ algorithm from the pertpy python library was used to identify cell clusters that credibly change in abundance with stimulation by either gRv or the MTB300 peptide pool compared to Media, relative to a reference cell type observed to remain stable across conditions. Only cell clusters observed to credibly increase with stimulation using this method were used for our analyses and the analysis was further restricted to cell cluster behavior in the two stimulation conditions, gRV and MTB300 peptide pool. Results from scCODA were confirmed by the use of Wilcoxon signed rank (paired) testing. In some cases, such as with downstream Mann-Whitney testing of abundance by progression status, the media condition cluster frequencies were subtracted from the stimulation cluster frequencies. This was done to account for the presence of background cell cluster activation. To account for clustering resolution bias, antigen-reactive cell clusters that exhibited similar phenotypes by gene markers were merged for qualitative and quantitative analyses.

Gene set enrichment analysis was used to identify differences between progressor and non-progressor groups within antigen-reactive cell types in the two stimulation conditions.

Stimulation conditions were analyzed separately. Raw integer counts were pseudobulked according to sample ID, stimulation condition, and annotated cell type with the AggregateExpression function from the Seurat R library ^62^. The edgeR ^63^ R library was used to identify metrics of differential gene expression, including logFC per gene with the Non-progressor group set as reference. Z score was calculated per gene using the zscoreT function from the limma ^64^ R library. Two different GSEA methods were used following edgeR: limma’s camera method ^65^ and fgsea ^66^. The Reactome gene set database ^67^ was used to search for enriched gene sets. Only gene sets found to be significant by both camera and fgsea were reported. The normalized enrichment scores (NES) from fgsea were used for visual representations of directionality of gene set enrichment. Samples were scored for each gene set found to be significant using the underlying scoring system from the score_terms function from the pathfindR ^68^ R library. Gene set leading edge genes by fgsea were used for scoring. The STRING protein-protein interaction database ^69^ was used to identify leading edge gene-gene relationships. STRING database values were parsed using the httr R library. The igraph ^70^, tidygraph, and ggplot2 R libraries were used for construction and visualization of the gene-gene networks.

## Data Availability

All data produced in the present study are available upon reasonable request to the authors.

## Supplmental Figure legends

**Supplemental Figure 1. scRNA-seq UMAP with ADT overlays**. A-F. UMAP of cells from 57 individuals labeled by major antigen-reactive cell lineages. A. CD3+ T cells. B. CD3+CD4+ T cells. C. CD3+CD8+ T cells. D. CD19+ B cells. E. CD56+ NK cells. F. CD11c monocytes and dendritic cells (mean denoised and scaled by background ADT counts per cluster^71^) .

**Supplemental Figure 2. scCODA identifies cell populations that significantly change in abundance in the presence of antigen stimulation.** A. CD4+ T cell cluster 1 is a CD4+ T central memory cell population that decreases in the presence of antigen. CD4+ T cell populations 9, 21, and 27 have different degrees of immune activation and cytokine transcript production and increase in the presence of antigen. B. NK cell population 10 is CD16+CD56+CD38+ and decreases in abundance in the presence of antigen. NK cell population 14 shows high expression of TNFRSF4 (OX40) and increases in the presence of antigen.

**Supplemental Figure 3. Abundance of antigen-reactive cell populations after in vitro stimulation in non-progressors and progressors.** A. Frequencies of MTB300-reactive monocytes (p=ns). B. Frequencies of gRv-reactive monocytes (p=ns). C. Frequencies of Mtb-reactive CD4+ T cells after stimulation with MTB300 or gRv (p=ns). D. Frequencies of Mtb-reactive NK cells after stimulation with MTB300 (p=ns), and after stimulation with gRv (higher in progressors, p=0.023).

## References

1. Lin, P.L. & Flynn, J.L. The End of the Binary Era: Revisiting the Spectrum of Tuberculosis. J Immunol 201, 2541–2548 (2018).

2. Sia, J.K. & Rengarajan, J. Immunology of Mycobacterium tuberculosis Infections. Microbiol Spectr 7 (2019).

3. Harari, A. et al. Dominant TNF-alpha+ Mycobacterium tuberculosis-specific CD4+ T cell responses discriminate between latent infection and active disease. Nature medicine 17, 372–376 (2011).

4. Mayer-Barber, K.D. & Barber, D.L. Innate and Adaptive Cellular Immune Responses to Mycobacterium tuberculosis Infection. Cold Spring Harbor perspectives in medicine 5 (2015).

5. Ernst, J.D. Mechanisms of M. tuberculosis Immune Evasion as Challenges to TB Vaccine Design. Cell Host Microbe 24, 34–42 (2018).

6. Vankayalapati, R. et al. Role of NK cell-activating receptors and their ligands in the lysis of mononuclear phagocytes infected with an intracellular bacterium. J Immunol 175, 4611–4617 (2005).

7. Dhiman, R. et al. IL-22 produced by human NK cells inhibits growth of Mycobacterium tuberculosis by enhancing phagolysosomal fusion. J Immunol 183, 6639–6645 (2009).

8. Schierloh, P. et al. NK cell activity in tuberculosis is associated with impaired CD11a and ICAM-1 expression: a regulatory role of monocytes in NK activation. Immunology 116, 541–552 (2005).

9. Venkatasubramanian, S. et al. IL-21-dependent expansion of memory-like NK cells enhances protective immune responses against Mycobacterium tuberculosis. Mucosal Immunol 10, 1031–1042 (2017).

10. Koeken, V., Verrall, A.J., Netea, M.G., Hill, P.C. & van Crevel, R. Trained innate immunity and resistance to Mycobacterium tuberculosis infection. Clin Microbiol Infect 25, 1468–1472 (2019).

11. Daley, C.L. et al. An outbreak of tuberculosis with accelerated progression among persons infected with the human immunodeficiency virus. An analysis using restriction-fragment-length polymorphisms. The New England journal of medicine 326, 231–235 (1992).

12. Keane, J. et al. Tuberculosis associated with infliximab, a tumor necrosis factor alpha-neutralizing agent. The New England journal of medicine 345, 1098–1104 (2001).

13. Mazurek, G.H. et al. Updated guidelines for using Interferon Gamma Release Assays to detect Mycobacterium tuberculosis infection - United States, 2010. MMWR Recomm Rep **59**, 1-25 (2010).

14. Barreto-Duarte, B. et al. Increased Frequency of Memory CD4+ T-Cell Responses in Individuals With Previously Treated Extrapulmonary Tuberculosis. Frontiers in immunology 11 (2020).

15. Covert, B.A., Spencer, J.S., Orme, I.M. & Belisle, J.T. The application of proteomics in defining the T cell antigens of Mycobacterium tuberculosis. Proteomics 1, 574–586 (2001).

16. Boesen, H., Jensen, B.N., Wilcke, T. & Andersen, P. Human T-cell responses to secreted antigen fractions of Mycobacterium tuberculosis. Infection and immunity 63, 1491–1497 (1995).

17. Arlehamn, C.S. et al. Dissecting mechanisms of immunodominance to the common tuberculosis antigens ESAT-6, CFP10, Rv2031c (hspX), Rv2654c (TB7.7), and Rv1038c (EsxJ). J Immunol **188**, 5020-5031 (2012).

18. Lein, A.D. et al. Cellular immune responses to ESAT-6 discriminate between patients with pulmonary disease due to Mycobacterium avium complex and those with pulmonary disease due to Mycobacterium tuberculosis. Clin Diagn Lab Immunol 6, 606–609 (1999).

19. Ravn, P. et al. Human T cell responses to the ESAT-6 antigen from Mycobacterium tuberculosis. The Journal of infectious diseases 179, 637–645 (1999).

20. Lalvani, A. et al. Human cytolytic and interferon gamma-secreting CD8+ T lymphocytes specific for Mycobacterium tuberculosis. Proceedings of the National Academy of Sciences of the United States of America 95, 270–275 (1998).

21. Lalvani, A. et al. Enumeration of T cells specific for RD1-encoded antigens suggests a high prevalence of latent Mycobacterium tuberculosis infection in healthy urban Indians. The Journal of infectious diseases 183, 469–477 (2001).

22. Lalvani, A. et al. Rapid detection of Mycobacterium tuberculosis infection by enumeration of antigen-specific T cells. American journal of respiratory and critical care medicine 163, 824–828 (2001).

23. Pathan, A.A. et al. Direct ex vivo analysis of antigen-specific IFN-gamma-secreting CD4 T cells in Mycobacterium tuberculosis-infected individuals: associations with clinical disease state and effect of treatment. J Immunol 167, 5217–5225 (2001).

24. Ubolyam, S. et al. Performance of a simple flow cytometric assay in diagnosing active tuberculosis. Tuberculosis 126, 102017 (2021).

25. Sauzullo, I. et al. Diagnostic performance in active TB of QFT-Plus assay and co-expression of CD25/CD134 in response to new antigens of Mycobacterium tuberculosis. Med Microbiol Immunol 208, 171–183 (2019).

26. Arlehamn, C.L. et al. Transcriptional profile of tuberculosis antigen-specific T cells reveals novel multifunctional features. J Immunol 193, 2931–2940 (2014).

27. Cai, Y. et al. Single-cell transcriptomics of blood reveals a natural killer cell subset depletion in tuberculosis. EBioMedicine 53, 102686 (2020).

28. Lindestam Arlehamn, C.S., et al. A Quantitative Analysis of Complexity of Human Pathogen-Specific CD4 T Cell Responses in Healthy M. tuberculosis Infected South Africans. PLoS pathogens 12, e1005760 (2016).

29. Arriaga, M.B. et al. Novel stepwise approach to assess representativeness of a large multicenter observational cohort of tuberculosis patients: The example of RePORT Brazil. Int J Infect Dis 103, 110–118 (2021).

30. Buttner, M., Ostner, J., Muller, C.L., Theis, F.J. & Schubert, B. scCODA is a Bayesian model for compositional single-cell data analysis. Nature communications 12, 6876 (2021).

31. Vankayalapati, R. et al. NK cells regulate CD8+ T cell effector function in response to an intracellular pathogen. J Immunol 172, 130–137 (2004).

32. Zhang, R., Zheng, X., Li, B., Wei, H. & Tian, Z. Human NK cells positively regulate gammadelta T cells in response to Mycobacterium tuberculosis. J Immunol 176, 2610–2616 (2006).

33. Roy, S. et al. NK cells lyse T regulatory cells that expand in response to an intracellular pathogen. J Immunol 180, 1729–1736 (2008).

34. Yu, J. et al. CD94 surface density identifies a functional intermediary between the CD56bright and CD56dim human NK-cell subsets. Blood 115, 274–281 (2010).

35. Chen, Z. et al. Phosphodiesterase 4A confers resistance to PGE2-mediated suppression in CD25(+) /CD54(+) NK cells. EMBO Rep 22, e51329 (2021).

36. Mao, Y. et al. IL-15 activates mTOR and primes stress-activated gene expression leading to prolonged antitumor capacity of NK cells. Blood 128, 1475–1489 (2016).

37. Crotty, S. A brief history of T cell help to B cells. Nature reviews. Immunology 15, 185–189 (2015).

38. Lindestam Arlehamn, C.S., et al. Memory T cells in latent Mycobacterium tuberculosis infection are directed against three antigenic islands and largely contained in a CXCR3+CCR6+ Th1 subset. PLoS pathogens 9, e1003130 (2013).

39. Acosta-Rodriguez, E.V. et al. Surface phenotype and antigenic specificity of human interleukin 17-producing T helper memory cells. Nature immunology 8, 639–646 (2007).

40. Perreau, M. et al. Lack of Mycobacterium tuberculosis-specific interleukin-17A-producing CD4+ T cells in active disease. European journal of immunology 43, 939–948 (2013).

41. Smith, C.A., Farrah, T. & Goodwin, R.G. The TNF receptor superfamily of cellular and viral proteins: activation, costimulation, and death. Cell 76, 959–962 (1994).

42. Locksley, R.M., Killeen, N. & Lenardo, M.J. The TNF and TNF receptor superfamilies: integrating mammalian biology. Cell 104, 487–501 (2001).

43. Ehlers, S. et al. The lymphotoxin beta receptor is critically involved in controlling infections with the intracellular pathogens Mycobacterium tuberculosis and Listeria monocytogenes. J Immunol 170, 5210–5218 (2003).

44. Netea, M.G. et al. Mycobacterium tuberculosis induces interleukin-32 production through a caspase-1/IL-18/interferon-gamma-dependent mechanism. PLoS medicine 3, e277 (2006).

45. Kim, S.H., Han, S.Y., Azam, T., Yoon, D.Y. & Dinarello, C.A. Interleukin-32: a cytokine and inducer of TNFalpha. Immunity 22, 131–142 (2005).

46. Netea, M.G. et al. Interleukin-32 induces the differentiation of monocytes into macrophage-like cells. Proceedings of the National Academy of Sciences of the United States of America 105, 3515–3520 (2008).

47. Netea, M.G. et al. Trained immunity: A program of innate immune memory in health and disease. Science 352, aaf1098 (2016).

48. Khader, S.A. et al. Targeting innate immunity for tuberculosis vaccination. The Journal of clinical investigation 129, 3482–3491 (2019).

49. Bailin, S.S. et al. Changes in subcutaneous white adipose tissue cellular composition and molecular programs underlie glucose intolerance in persons with HIV. Frontiers in immunology 14, 1152003 (2023).

50. Zheng, G.X. et al. Massively parallel digital transcriptional profiling of single cells. Nature communications 8, 14049 (2017).

51. Alves, E. et al. Underrepresentation of activating KIR gene expression in single-cell RNA-seq data is due to KIR gene misassignment. European journal of immunology, e2350590 (2023).

52. Heaton, H. et al. Souporcell: robust clustering of single-cell RNA-seq data by genotype without reference genotypes. Nature methods 17, 615–620 (2020).

53. Dill-McFarland, K.A. et al. Genome-wide association study in Brazil identifies genetic susceptibility to tuberculosis with single-cell gene expression effects. medRxiv (2025).

54. Neavin, D. et al. Demuxafy: improvement in droplet assignment by integrating multiple single-cell demultiplexing and doublet detection methods. Genome Biol 25, 94 (2024).

55. Bernstein, N.J. et al. Solo: Doublet Identification in Single-Cell RNA-Seq via Semi-Supervised Deep Learning. Cell Systems 11, 95–101.e105 (2020).

56. Gayoso, A. et al. A Python library for probabilistic analysis of single-cell omics data. Nature biotechnology 40, 163–166 (2022).

57. Virshup, I. et al. The scverse project provides a computational ecosystem for single-cell omics data analysis. Nature biotechnology 41, 604–606 (2023).

58. Linderman, G.C., Zhao, J. & Kluger, Y. Zero-preserving imputation of scRNA-seq data using low-rank approximation. bioRxiv, 397588 (2018).

59. Korsunsky, I. et al. Fast, sensitive and accurate integration of single-cell data with Harmony. Nature methods 16, 1289–1296 (2019).

60. Traag, V.A., Waltman, L. & van Eck, N.J. From Louvain to Leiden: guaranteeing well-connected communities. Scientific reports 9, 5233 (2019).

61. Healy, J. & McInnes, L. Uniform manifold approximation and projection. Nature Reviews Methods Primers 4, 82 (2024).

62. Hao, Y. et al. Dictionary learning for integrative, multimodal and scalable single-cell analysis. Nature biotechnology 42, 293–304 (2024).

63. Chen, Y., Chen, L., Lun, Aaron T.L., Baldoni, Pedro L. & Smyth, Gordon K. edgeR v4: powerful differential analysis of sequencing data with expanded functionality and improved support for small counts and larger datasets. Nucleic Acids Research 53 (2025).

64. Ritchie, M.E. et al. limma powers differential expression analyses for RNA-sequencing and microarray studies. Nucleic Acids Research 43, e47–e47 (2015).

65. Wu, D. & Smyth, G.K. Camera: a competitive gene set test accounting for inter-gene correlation. Nucleic Acids Research 40, e133–e133 (2012).

66. Korotkevich, G. et al. Fast gene set enrichment analysis. bioRxiv, 060012 (2021).

67. Milacic, M. et al. The Reactome Pathway Knowledgebase 2024. Nucleic Acids Research 52, D672-D678 (2023).

68. Ulgen, E., Ozisik, O. & Sezerman, O.U. pathfindR: An R Package for Comprehensive Identification of Enriched Pathways in Omics Data Through Active Subnetworks. Front Genet 10, 858 (2019).

69. Szklarczyk, D. et al. The STRING database in 2023: protein–protein association networks and functional enrichment analyses for any sequenced genome of interest. Nucleic Acids Research 51, D638–D646 (2022).

70. Csárdi, G. et al. igraph for R: R interface of the igraph library for graph theory and network analysis. 2024 [cited]Available from: 10.5281/zenodo.10602726

71. Mule, M.P., Martins, A.J. & Tsang, J.S. Normalizing and denoising protein expression data from droplet-based single cell profiling. Nature communications 13, 2099 (2022).

