## Supplementary figures and images for "Single-cell immune profiling at time of *M. tuberculosis* exposure reveals antigen-reactive programs that predict progression to active disease"

### Supplemental Figures 1-3

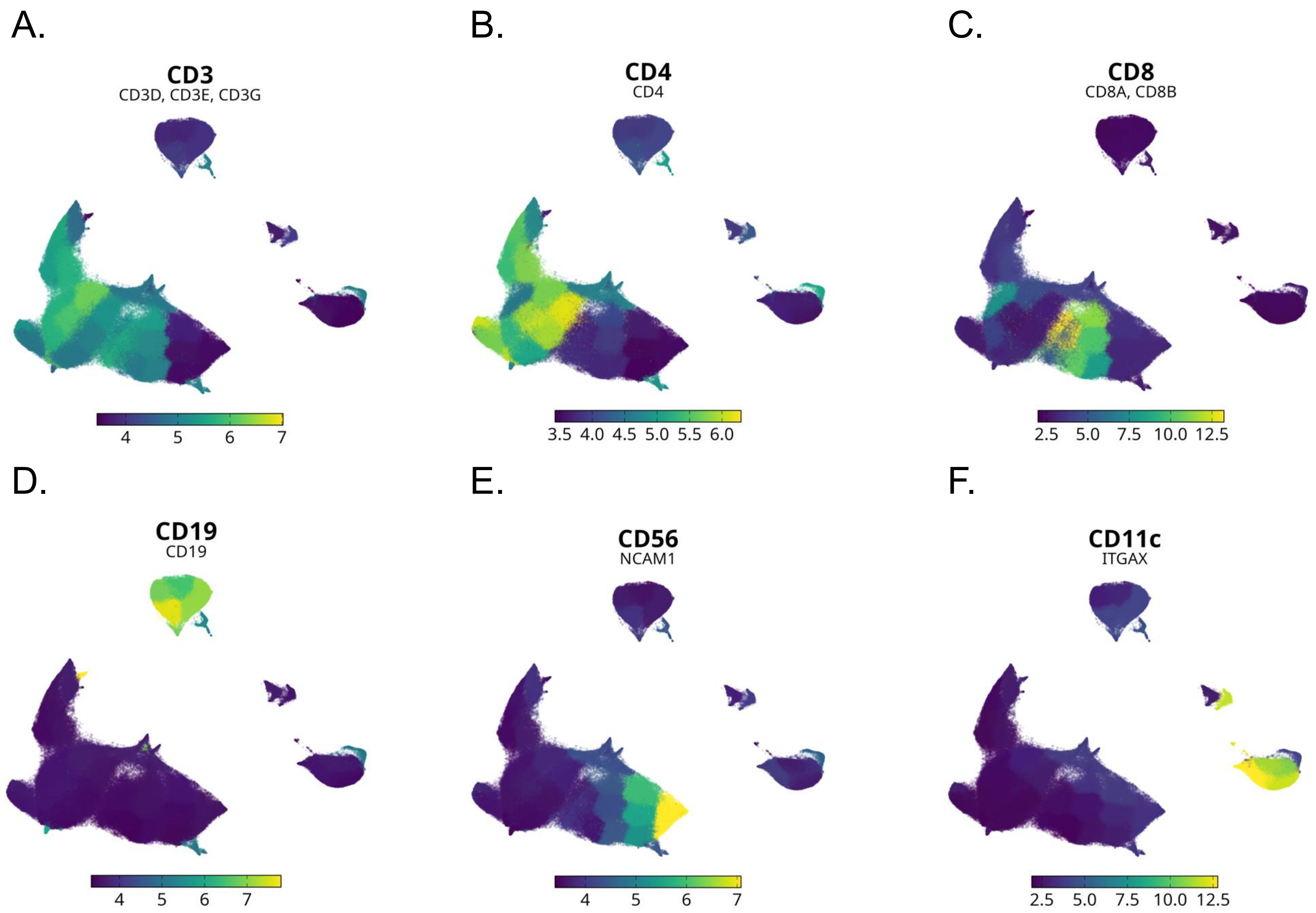

A. CD4+ T cell populations

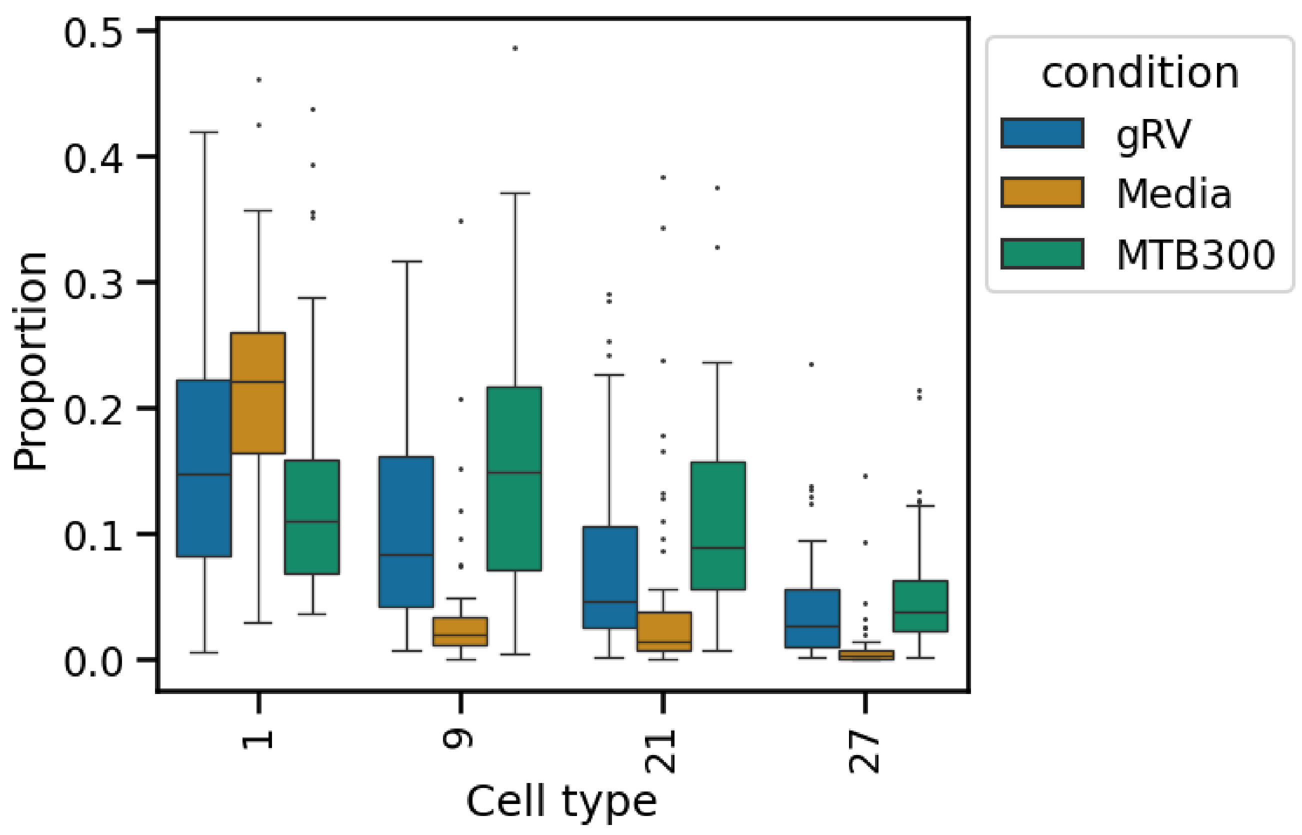


B. NK cell populations

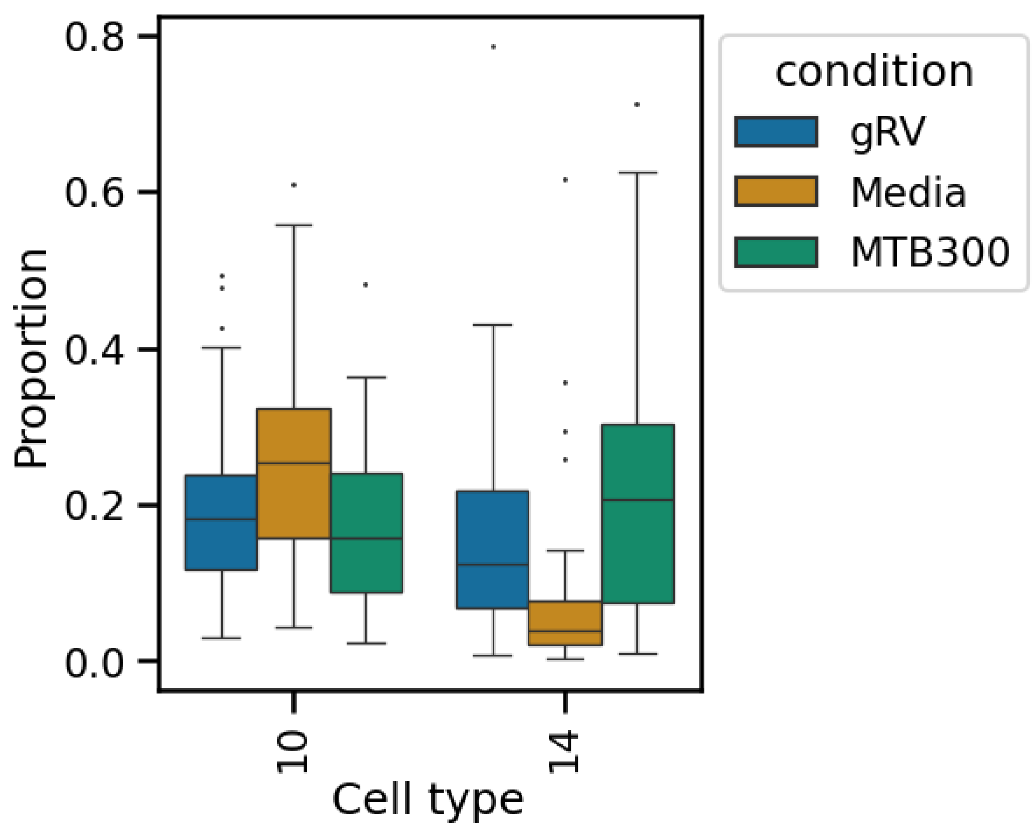

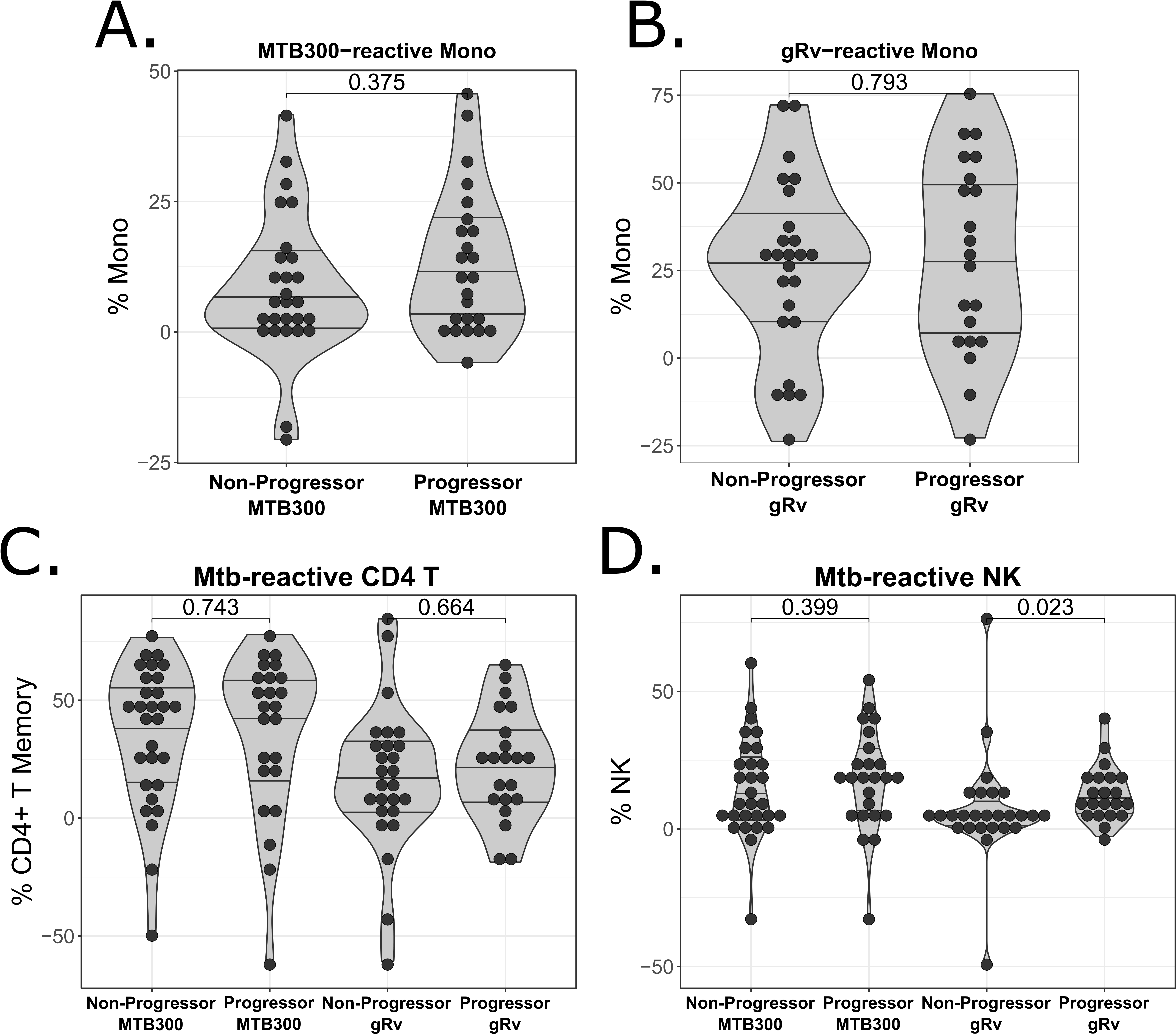
